# Association of HLA class I genotypes with age at death of COVID-19 patients

**DOI:** 10.1101/2020.11.19.20234567

**Authors:** Maxim Shkurnikov, Stepan Nersisyan, Tatjana Jankevic, Alexei Galatenko, Ivan Gordeev, Valery Vechorko, Alexander Tonevitsky

**Affiliations:** Faculty of Biology and Biotechnology, HSE University, Moscow, Russia; Center for Precision Genome Editing and Genetic Technologies for Biomedicine, Pirogov Russian National Research Medical University, Moscow, Russia; Faculty of Mechanics and Mathematics, Lomonosov Moscow State University, Moscow, Russia; O.M. Filatov City Clinical Hospital, Moscow, Russia; Shemyakin-Ovchinnikov Institute of Bioorganic Chemistry, Russian Academy of Sciences, Moscow, Russia

## Abstract

**Background:** HLA class I molecules play a crucial role in the development of a specific immune response to viral infections by presenting viral peptides to cell surface where they will be further recognized by T cells. In the present manuscript we explored whether HLA class I genotype can be associated with critical course of COVID-19 by searching possible connections between genotypes of deceased patients and their age at death.

**Methods and Findings:** HLA-A, HLA-B and HLA-C genotypes of *n* = 111 deceased patients with COVID-19 (Moscow, Russia) and *n* = 428 volunteers were identified with targeted next-generation sequencing. Deceased patients were splitted into two groups according to age at death: *n* = 26 adult patients with age at death below 60 completed years (inclusively) and *n* = 85 elderly patients over 60. With the use of HLA class I genotypes we developed a risk score which is associated with the ability to present SARS-CoV-2 peptides by an individual’s HLA class I molecule set. The resulting risk score was significantly higher in the group of deceased adults compared to elderly adults (*p* = 0.00348, AUC ROC = 0.68). In particular, presence of HLA-A*01:01 allele was associated with high risk, while HLA-A*02:01 and HLA-A*03:01 mainly contributed to the low risk group. The analysis of homozygous patients highlighted the results even stronger: homozygosity by HLA-A*01:01 mainly accompanied early deaths, while only one HLA-A*02:01 homozygote died before 60.

**Conclusions:** The obtained results suggest the important role of HLA class I peptide presentation in the development of a specific immune response to COVID-19. While prediction of age at death by HLA class I genotype had a reliable performance, involvement of HLA class II genotype can make it even higher in the future studies.

## Introduction

HLA class I molecules are one of the key mediators of the first links in the development of a specific immune response to COVID-19 infection. Right after entering the cell SARS-CoV-2 induces the translation of its proteins. Some of these proteins enter the proteasomes of the infected cell, get cleaved to peptides of the length 8-12 amino acid residues and bind to HLA class I receptors. After binding the complex consisting of the HLA class I molecule and the peptide is transferred to the surface of the infected cell, where it can interact with the T cell receptor of CD8+ T lymphocytes. In response to the interaction the CD8+ T lymphocyte activates and starts to divide; in 5-7 days a population of virus-specific cytotoxic CD8+ T lymphocytes capable of destroying infected cells using perforins and serine proteases gets formed [1].

There are three main types of HLA class I receptors: HLA-A, HLA-B and HLA-C. Receptors of every type are present in two variants inherited from parents. There exist dozens of variants of each allele of HLA-I receptors; every allele has an individual ability to recognize various foreign proteins. The distribution of alleles is population/country specific [2].

Individual combinations of HLA class I receptors essentially affect the severity of multiple infectious diseases, including malaria [3], tuberculosis [4], HIV [5] and viral hepatitis [2]. There is a number of reported interconnections between the HLA genotype and sensitivity to SARS-CoV. E.g. the alleles HLA-B*07:03 [6], HLA-B*46:01 [7] and HLA-C*08:01 [8] are factors of predisposition to a severe form of the disease; the allele HLA-C*15:02 is associated with a mild form [9].

Information on the interconnection of HLA class I genotype and severity of the course of the new coronavirus infection (COVID-19) caused by SARS-CoV-2 is sparse. A sample of 45 patients with varying severity of COVID-19 was used to confirm the results of theoretical modeling of interaction of SARS-CoV-2 peptides with various HLA-I alleles [10]. It was demonstrated that the number of peptides with a high interaction constant is connected with individual HLA genotype: the more viral peptides with high affinity bind to HLA class I, the easier is the course of the disease. It was also shown that the frequency of the occurrence of HLA-A*01:01 and HLA-A*02:01 alleles is related to the number of infections and mortality rate in different regions of Italy [11].

In the present study we explored whether HLA class I genotype can be a factor contributing to the critical course of COVID-19. For that we performed HLA genotyping for *n* = 111 deceased patients with COVID-19 as well as the control group (*n* = 428), and searched for putative associations between genotypes and age at death. Since the total number of distinct HLA class I genotypes is too high for performing frequency-based analysis, we assigned scores to each allele based on capability of presenting SARS-CoV-2 peptides. The obtained scores allowed us to make valid statistical comparison of HLA genotypes in groups of deceased adults (age at death not greater than 60 completed years, *n* = 26), elderly adults (age at death over 60, *n* = 85) and the control. Special attention was paid to “extreme” cases formed by individuals homozygous by some of HLA genes. Additionally, we assessed the contribution of each viral protein to the constructed risk model.

## Materials and methods

### Design and participants

There were 111 patients infected with COVID-19 enrolled in O.M. Filatov City Clinical Hospital, (Moscow, Russia) who died between May and July 2020. All patients had at least one positive test result for SARS-CoV-2 by RT-qPCR from nasopharyngeal swabs or bronchoalveolar lavage. Patients with pathologies that lead to greater morbidity or who had additional immunosuppression (HIV, active cancer in treatment with chemotherapy, immunodeficiency, autoimmune diseases with immunosuppressants, transplant patient) were not included in the study. Blood (2 ml) was collected by the medical practitioner from the right ventricle in an EDTA vial *post-mortem*. Patients were divided into two groups according to their age of death: adults (age ≤ 60, *n* = 26) and elderly adults (age *>* 60, *n* = 85).

The control group of 428 volunteers was established with the use of electronic HLA genotype records of the Federal register of bone marrow donors (Pirogov Russian National Research Medical University). All patients or their next of kin gave informed consent for participation in the study.

The study protocol was reviewed and approved by the Local Ethics Committee at the Pirogov Russian National Research Medical University (Meeting No. 194 of March 16 2020, Protocol No. 2020/07); the study was conducted in accordance with the Declaration of Helsinki.

### HLA class I genotyping with targeted next-generation sequencing

Genomic DNA was isolated from frozen-collected anticoagulated whole blood samples by using the QIAamp DNA Blood Mini Kit on the automatic workstation QIACube (QIAGEN GmbH, Hilden, Germany). HLA-A, HLA-B and HLA-C genes were sequenced with MiSeq platform (Illumina, San Diego, CA, USA) through exons 2 to 4 in both directions using reagent kit HLA-Expert (DNA-Technology LLC, Moscow, Russia) and annotated using the database of the human major histocompatibility complex sequences IMGT/HLA v3.41.0 [12]. Processed genotype data is available in S1 Table.

### SARS-CoV-2 protein sequences

Publicly available SARS-CoV-2 proteomes derived from patients infected in Moscow (*n* = 79) were obtained from GISAID [13] (full list of IDs is presented within S2 Table). Clustal Omega v1.2.4 was used to construct multiple sequence alignment for each viral protein [14]. The obtained alignment had no gaps and rare mutations: only 117 out of 9719 positions (1.2%) contained more than one amino acid variant. Moreover, distribution of non-major amino acid fractions at mutation sites was also concentrated near zero: maximum fraction was equal to 22.8% (18 out of 79 viruses), 0.95 quantile was equal to 5.1% (4 viruses) and upper quartile was equal to 1.3% (one virus with mismatched amino acid).

Since many research groups use the reference SARS-CoV-2 genome and proteome sequences (Wuhan-Hu-1, NCBI Reference Sequence: NC 045512.2) we compared the obtained alignment with the mentioned reference. Two protein sequences differed in four positions (0.04%) located within NSP12, N and S viral proteins. Such differences can indeed lead to functional diversity of analyzed proteins, however will be totally negligible for the further analysis.

### Prediction of viral peptides and assessment of their binding affinities to HLA class I molecules

We applied the procedure described by Nguen et al in [15] to the consensus protein sequences of viruses isolated from patients in Moscow. Specifically, for each amino acid of each viral protein we assessed the probability of proteasomal cleavage in the considered site using NetChop v3.1 [16]. The set of viral peptides was generated by taking all possible 8-mers to 12-mers having proteasomal cleavage probability not less than 0.1 at both ends of a sequence.

Binding affinities were predicted using netMHCpan v4.1 [17] for all viral peptides (*n* = 15314) and HLA alleles present in our cohorts of deceased and control patients (*n* = 107). Peptides having weak binding affinity to all considered alleles were discarded (*IC*_50_ affinity values above 500 nM as recommended by netMHCpan developers). For the remaining 6548 peptides all affinities were inverted, multiplied by 500 and log_10_-transformed. Thus, the resulting score was equal to zero for peptides with weak binding affinity threshold (500 nM) and equal to one for the high binding affinity (50 nM). Raw and processed matrices are presented in S3 Table.

### Statistical analysis

Allele frequencies in considered cohorts were estimated by dividing the number of occurrences of a given allele in individuals by the doubled total number of individuals (i.e. identical alleles of homozygous individuals were counted as two occurences). The following functions from scipy.stats Python module [18] were used to conduct statistical testing: fisher exact for Fisher’s exact test, mannwhitneyu for Mann-Whitney U test. Benjamini-Hochberg procedure was used to perform multiple testing correction. Principal component analysis was conducted with scikit-learn Python module [19]. Permutation test for assessing significance of area under the receiver operating characteristic curve (AUC ROC) values was done with *n* = 10^6^ label permutations. Plots were constructed with Seaborn and Matplotlib [20].

## Results

### Distribution of HLA class I gene alleles in the cohort of deceased COVID-19 patients and the control group

We performed HLA class I genotyping for *n* = 111 deceased patients with confirmed COVID-19 (Moscow, Russia) and the control group consisting of volunteers (*n* = 428). Deceased patients were divided into two groups: adults (age at death less or equal to 60 years) and elderly adults (age at death over 60 years). Demographic and clinical data of these cohorts is summarized in Table 1. Although patients with severe comorbidities were excluded from the study, 76.6 % of deceased patients had at least one underlying disease. Only cerebrovascular disease had statistically significant odds ratio when comparing groups of adults and elderly adults (3.8% versus 34.1%, Fisher’s exact test *p* = 1.89× 10^−3^). Other cardiovascular diseases like coronary artery disease and heart failure were also more frequent in group of elders which, however, was not statistically significant. Interestingly, arterial hypertension was diagnosed in 11.5% adult patients and 24.7% older adults which was generally less than populational level in Russia (about 50%) [21]. Percentage of diabete cases was about 3.5% in both analyzed groups which is a typical value for the current population of Russia [22]. Also, frequencies of chronic kidney disease (stages 4–5) in both groups (23.1% for adults and 16.5% for elders) was significantly higher compared to background populational value (about 0.05%) [23].

**Table 1.** Demographic and clinical data in the cohort of deceased patients with COVID-19.

| | Deceased,<br>age $\leq 60$,<br>$n = 26$ | Deceased,<br>age $> 60$,<br>$n = 85$ | $p$ |
| --- | --- | --- | --- |
| Age, completed years,<br>median (Q25–Q75) | 54.5 (45.25–57) | 79 (70–84) |  |
| **Gender** |  |  |  |
| Female | 10 (38.5%) | 42 (49.4%) | 0.375 |
| Male | 16 (61.5%) | 43 (50.6%) | 0.375 |
| **Comorbidities** |  |  |  |
| None | 8 (30.8%) | 18 (21.2%) | 0.305 |
| 1 | 11 (42.3%) | 43 (50.6%) | 0.507 |
| $> 1$ | 7 (26.9%) | 24 (28.2%) | 1.000 |
| **Cardiovascular disease** |  |  |  |
| Arterial Hypertension | 3 (11.5%) | 21 (24.7%) | 0.184 |
| Coronary artery disease | 1 (3.8%) | 9 (10.6%) | 0.448 |
| Cardiac infarction | 1 (3.8%) | 2 (2.4%) | 0.555 |
| Heart failure | 0 (0.0%) | 9 (10.6%) | 0.113 |
| Stroke | 2 (7.7%) | 7 (8.2%) | 1.000 |
| Cerebrovascular disease | 1 (3.8%) | 29 (34.1%) | $1.89 \times 10^{-3}$ |
| **Metabolic disease** |  |  |  |
| Obesity | 1 (3.8%) | 1 (1.2%) | 0.415 |
| Diabetes | 1 (3.8%) | 3 (3.5%) | 1.000 |
| Chronic obstructive<br>pulmonary disease | 2 (7.7%) | 3 (3.5%) | 0.333 |
| **Neoplasma** | 5 (19.2%) | 8 (9.4%) | 0.179 |
| **Others** |  |  |  |
| Alcoholic liver disease | 2 (7.7%) | 2 (2.4%) | 0.233 |
| Gastric ulcer | 3 (11.5%) | 3 (3.5%) | 0.140 |
| Chronic kidney disease,<br>stages 4–5 | 6 (23.1%) | 14 (16.5%) | 0.560 |

Comorbidity ratios were compared with Fisher’s exact test.

First, we tested whether frequency of a single allele can differentiate individuals from three groups: adult patients died from COVID-19, elderly patients died from COVID-19 and the control group. Distribution of major HLA-A, HLA-B and HLA-C alleles in these three groups is summarized in Fig 1. Fisher’s exact test was used to make formal statistical comparisons. As a result, we found that for all possible group comparisons not a single allele had odds ratio which can be considered statistically significant after multiple testing correction (all corrected *p*-values were equal to 1). However, few of them were differentially enriched if no multiple testing correction was applied (S4 Table).

**Fig 1.**
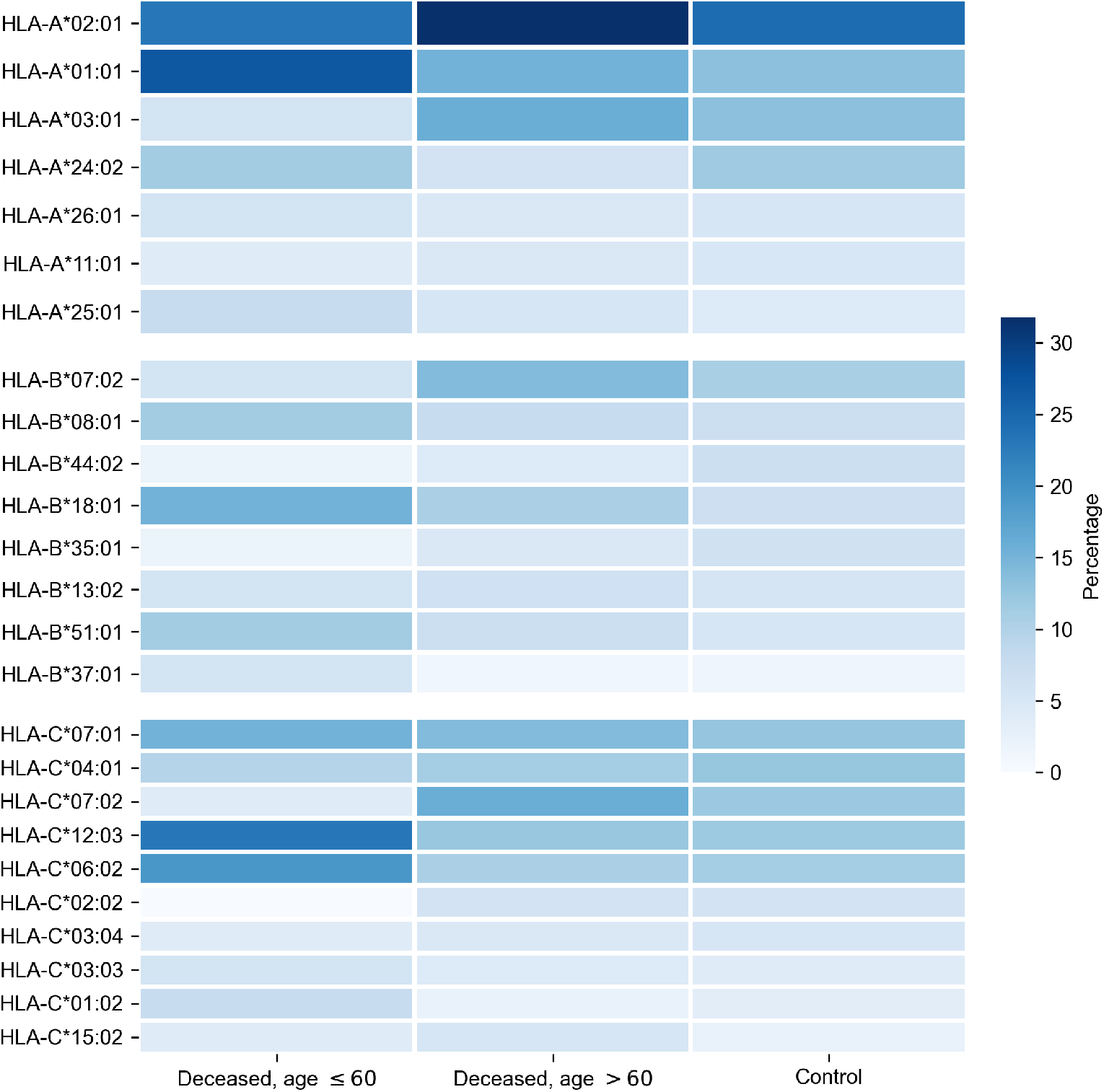
Distribution of HLA-A, HLA-B, HLA-C alleles in cohorts of deceased COVID-19 patients and the control group. Alleles with frequency over 5% in at least one of three considered groups are presented.

### Binding affinities of viral peptides to HLA class I molecules

Since sizes of considered cohorts were insufficient for performing frequency analysis at level of full HLA class I genotypes, we transformed patient genotypes from discrete space to numerical units associated with the potential of interactions with SARS-CoV-2 peptides. To implement this idea we first constructed a matrix of binding affinities of viral peptides to HLA-A, HLA-B and HLA-C alleles. For that we first made computational predictions of viral peptides derived from SARS-CoV-2 strains isolated in Moscow. Then, binding affinities were calculated for each of predicted peptides and each allele present in patients from analyzed cohorts.

As a result, we obtained the matrix containing affinity values for 6548 peptides and 107 alleles of genes from major HLA class I. To establish a positive relationship between values from matrix and binding potential, all affinities were inverted and scaled by value of 500 nM (the conventional threshold for binding ability). Simultaneous hierarchical clustering was applied to identify groups of similar peptides and HLA-A, HLA-B and HLA-C alleles (Fig 2). As can be seen, both alleles and peptides clearly formed several dense clusters. The most presented alleles of HLA-A (HLA-A*01:01, HLA-A*02:01, HLA-A*03:01, HLA-A*24:02) fell in different clusters, while for HLA-B and HLA-C some major alleles grouped together (e.g. HLA-B*07:02, HLA-B*08:01 and HLA-C*06:02, HLA-C*07:01, HLA-C*07:02).

**Fig 2.**
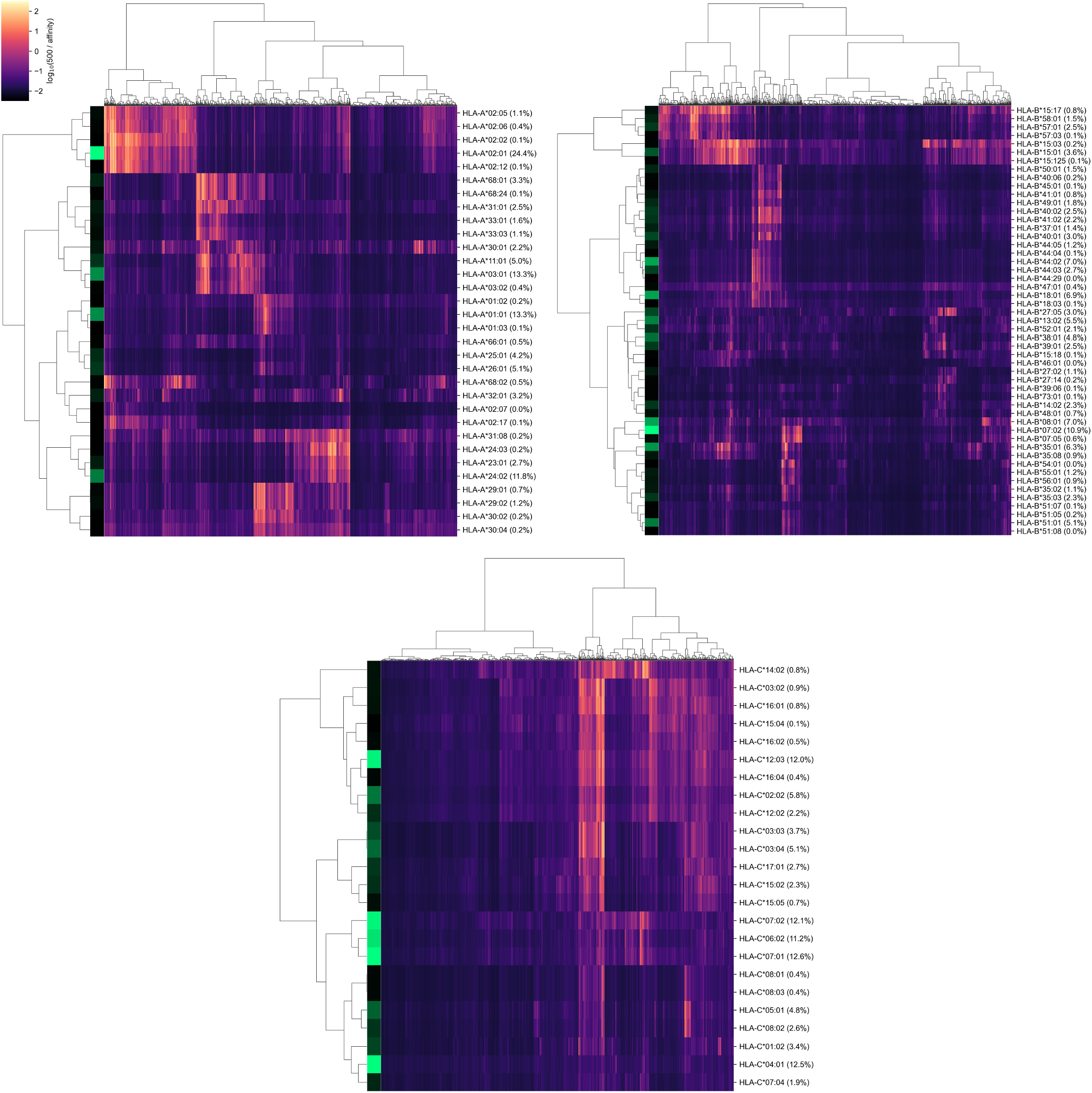
Hierarchical clustering of HLA-A, HLA-B, HLA-C gene alleles and SARS-CoV-2 peptides according to binding affinity matrix. Shades of green in vertical stripes and percents in brackets represent frequency of an allele in the control group. Zero percents refer to rare alleles found only in the group of deceased patients.

Note that alleles with similar peptide binding profiles can be linked to different alleles of remaining genes. For example, consider closely clustered alleles HLA-C*06:02, HLA-C*07:01 and HLA-C*07:02. From the analysis of the contingency table of allele pairs in the control group (Fig 3) it follows that each of these alleles has its own spectrum of associated alleles. Specifically, HLA-C*06:02 usually appears with HLA-B*13:02 (Fisher’s exact test *p* = 3.18 × 10^−14^), HLA-C*07:01 is linked to HLA-A*01:01 (*p* = 2.97 × 10^−7^) and HLA-B*08:01 (*p* = 3.15 × 10^−14^), while HLA-C*07:02 is coupled with HLA-A*03:01 (*p* = 9.66 × 10^−4^) and HLA-B*07:02 (*p* = 2.72 × 10^−26^). Interestingly, such linked alleles can have different peptide binding patterns (e.g. see weakly overlapped bars for HLA-A*01:01 and HLA-A*03:01 in Fig 2).

**Fig 3.**
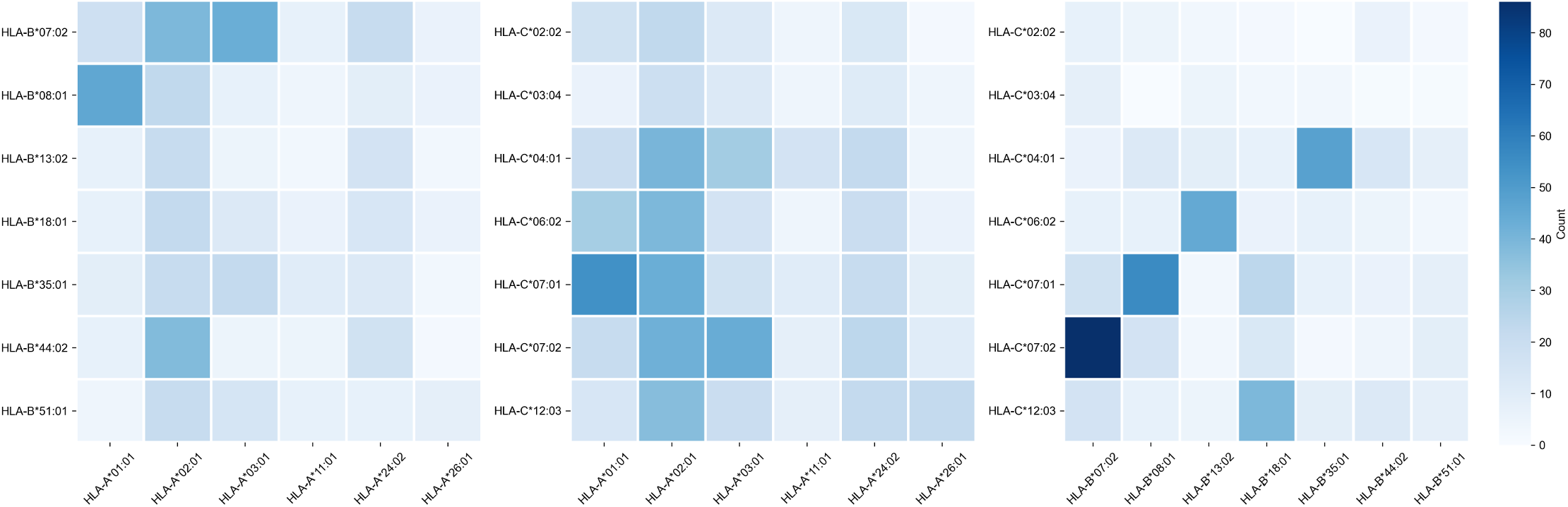
Contingency table of allele counts in the control group. Alleles with frequency over 5% in the control group are presented.

### Risk score based on peptide-HLA binding affinity is associated with early COVID-19 deaths

For each of considered HLA-A, HLA-B and HLA-C alleles we obtained the list of binding affinities to 6548 unique SARS-CoV-2 peptides. In order to calculate aggregate information on potential of presenting SARS-CoV-2 peptides by each of alleles, we used principal component analysis (PCA). In this framework 6548-element affinity vectors are replaced by the most informative linear combinations of their components. For HLA-A and HLA-C we found four principal components (PCs) each of which explained at least 5% of data variance, while for HLA-B the number of essential components was equal to five (S5 Table). Signs of components were set in the way to achieve positive correlation of component values with age of death of deceased patients.

For each individual, HLA class I gene and PC we summed PC values associated with two corresponding alleles. After that we analyzed differences of obtained scores in adult and elderly patients who died from COVID-19. Three of the resulting PCs demonstrated statistically significant differences according to Mann-Whitney U test. This list included the second and the third PCs of HLA-A, and the fourth principal component of HLA-C, while no PCs of HLA-B separated analyzed groups significantly. As an aggregate risk score (RS) we considered the sum of these three components (for convenience, we linearly scaled the range of RS to the [0, 100] interval). The obtained score also significantly separated groups with *p* = 3.48×10^−3^ (U test) and area under the receiver operating characteristic curve (AUC ROC) equal to 0.68 (permutation test *p* = 3.10×10^−3^), see Fig 4 (full information is listed in S6 Table). Interestingly, the difference of RS distributions in the cohort of adult patients and the control group was also statistically significant (U test *p* = 3.31×10^−3^), while the difference between elderly and control groups was not (*p* = 0.283).

**Fig 4.**
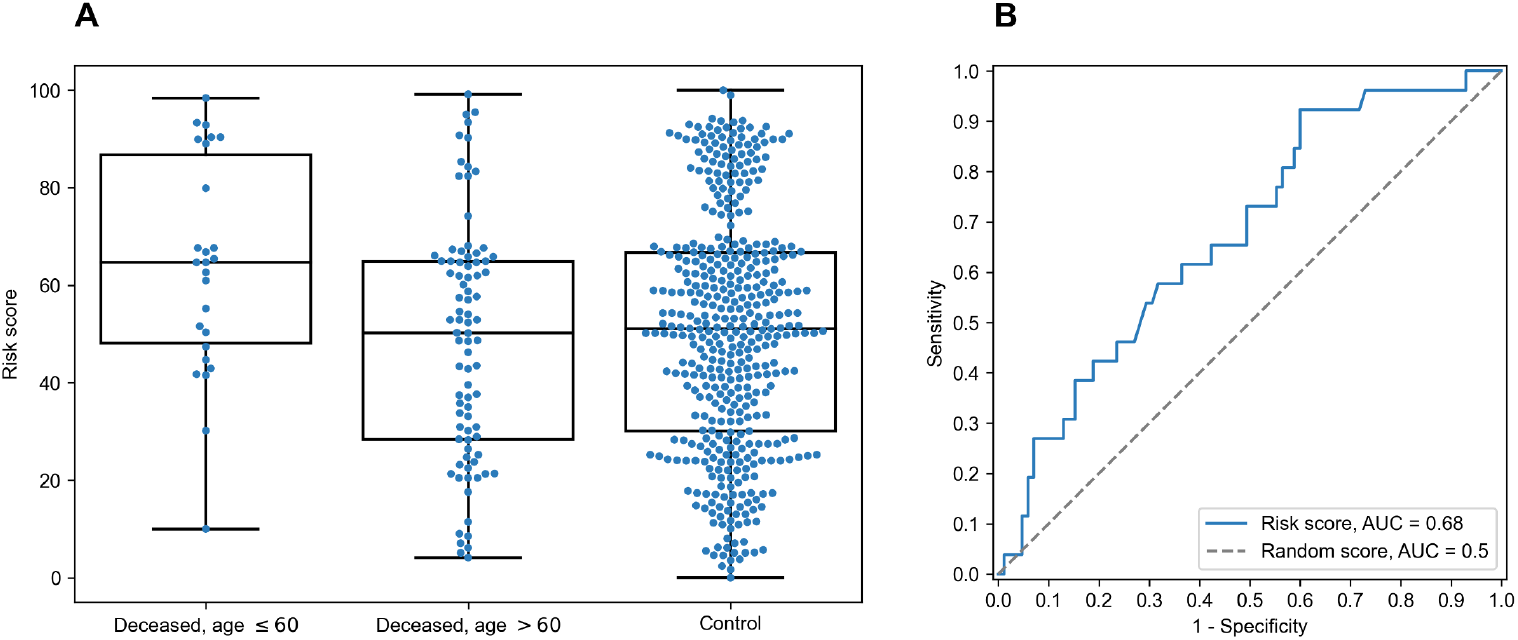
Risk score (RS) separates groups of adult and elderly patients. A: distribution of RS in adult, elderly and control patient groups. B: receiver operating characteristic curve for RS separating patients from adult and elderly groups.

In order to characterize the association between RS and age at death more precisely, we partitioned the range of RS into three groups: low, medium and high (Fig 5). The lower and higher thresholds were calculated in a way to minimize *p*-value for Fisher’s exact test applied to the number of adult and elderly patients in the whole cohort and in the low/high risk groups, respectively. Interestingly, such partitioning led to significant separation of adult patients both from elderly and control ones in low and high risk groups, while no significance was found within the middle group (S7 Table).

**Fig 5.**
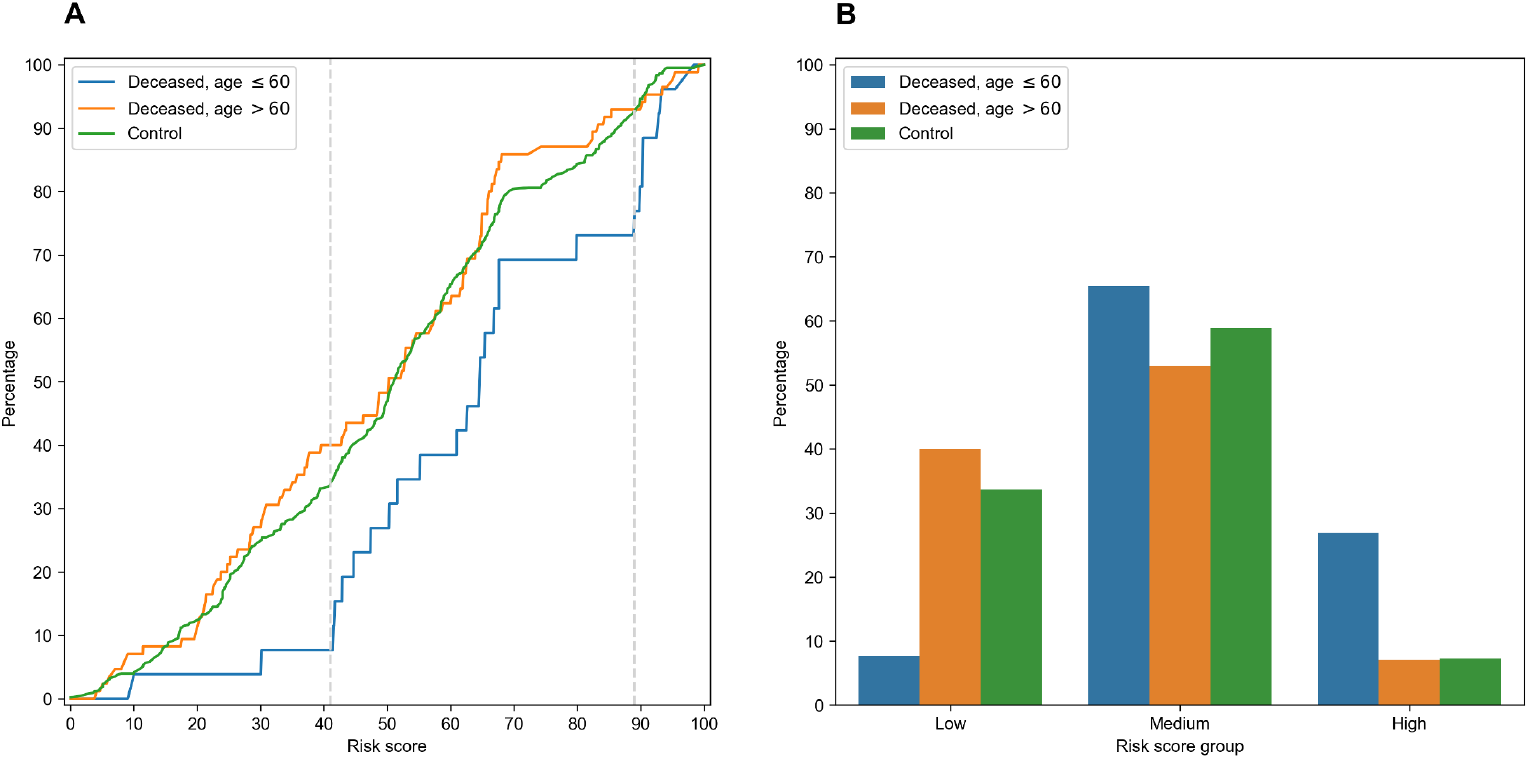
Low, middle and high risk score (RS) groups. A: empirical distribution function of RS in three patient groups. Vertical dotted lines at RS = 41 and RS = 89 define ranges for three RS groups. B: distribution of three patient groups over low, medium and high RS.

Then, we performed enrichment analysis to identify alleles significantly contributing to each of RS groups (Table 2). As it can be seen, frequencies of several alleles were dramatically higher in some of RS groups. Specifically, HLA-A*02:01 and HLA-A*03:01 were highly overrepresented in low risk and completely absent at high risk groups, while the most enriched allele in high risk group was HLA-A*01:01. Reciprocally to HLA-A*02:01 and HLA-A*03:01 cases, not a single individual in low risk group carried HLA-A*01:01 allele. Complete information on allele frequencies in RS groups is presented in S1 Fig.

**Table 2.**
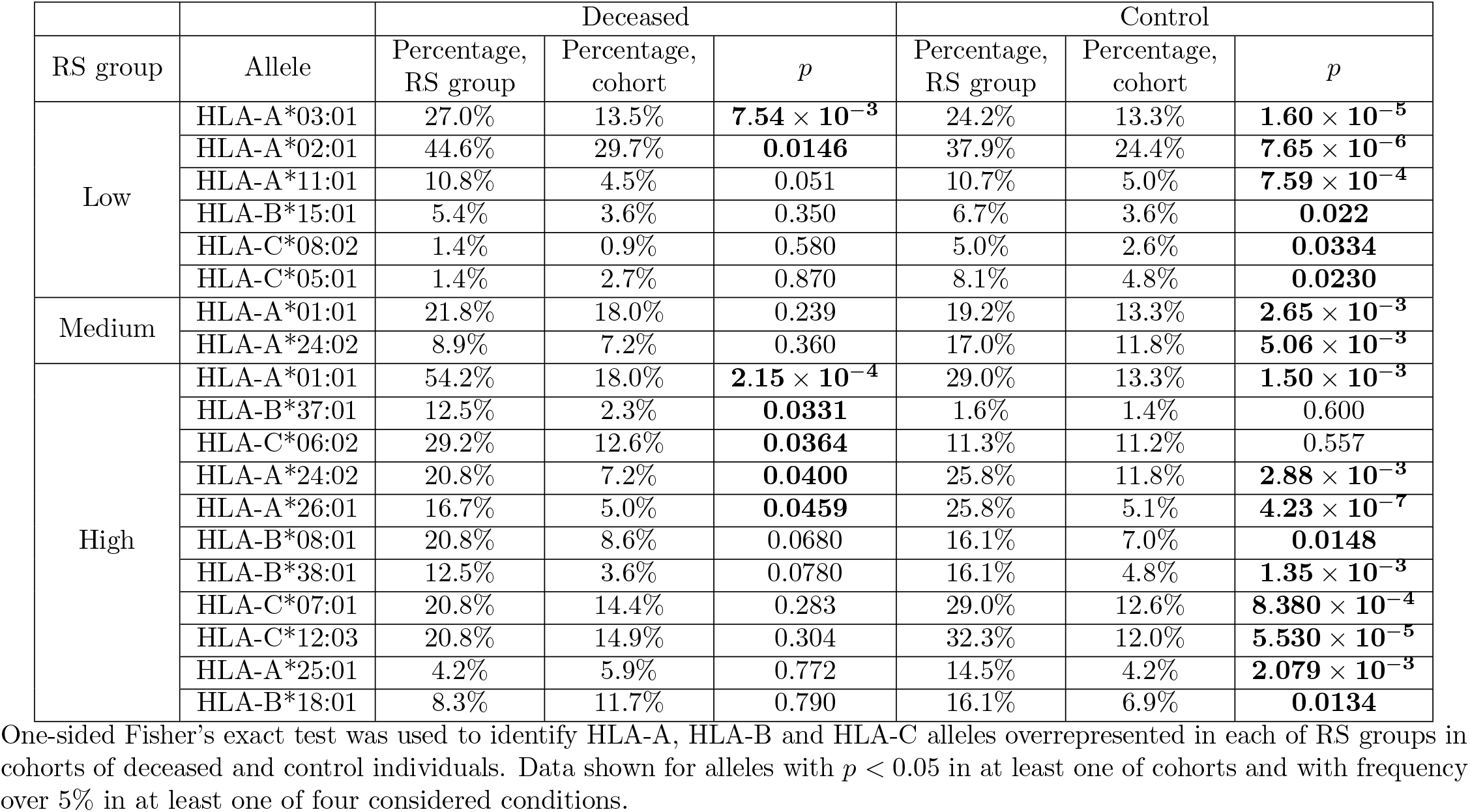
Enrichment analysis of risk score groups.

| RS group | Allele | Deceased |  |  | Control |  |  |
| --- | --- | --- | --- | --- | --- | --- | --- |
| | | Percentage, RS group | Percentage, cohort | $p$ | Percentage, RS group | Percentage, cohort | $p$ |
| Low | HLA-A*03:01 | 27.0% | 13.5% | $7.54 \times 10^{-3}$ | 24.2% | 13.3% | $1.60 \times 10^{-5}$ |
| | HLA-A*02:01 | 44.6% | 29.7% | **0.0146** | 37.9% | 24.4% | $7.65 \times 10^{-6}$ |
| | HLA-A*11:01 | 10.8% | 4.5% | 0.051 | 10.7% | 5.0% | $7.59 \times 10^{-4}$ |
|  | HLA-B*15:01 | 5.4% | 3.6% | 0.350 | 6.7% | 3.6% | **0.022** |
|  | HLA-C*08:02 | 1.4% | 0.9% | 0.580 | 5.0% | 2.6% | **0.0334** |
|  | HLA-C*05:01 | 1.4% | 2.7% | 0.870 | 8.1% | 4.8% | **0.0230** |
| Medium | HLA-A*01:01 | 21.8% | 18.0% | 0.239 | 19.2% | 13.3% | $2.65 \times 10^{-3}$ |
| | HLA-A*24:02 | 8.9% | 7.2% | 0.360 | 17.0% | 11.8% | $5.06 \times 10^{-3}$ |
| High | HLA-A*01:01 | 54.2% | 18.0% | $2.15 \times 10^{-4}$ | 29.0% | 13.3% | $1.50 \times 10^{-3}$ |
|  | HLA-B*37:01 | 12.5% | 2.3% | **0.0331** | 1.6% | 1.4% | 0.600 |
|  | HLA-C*06:02 | 29.2% | 12.6% | **0.0364** | 11.3% | 11.2% | 0.557 |
| | HLA-A*24:02 | 20.8% | 7.2% | **0.0400** | 25.8% | 11.8% | $2.88 \times 10^{-3}$ |
| | HLA-A*26:01 | 16.7% | 5.0% | **0.0459** | 25.8% | 5.1% | $4.23 \times 10^{-7}$ |
|  | HLA-B*08:01 | 20.8% | 8.6% | 0.0680 | 16.1% | 7.0% | **0.0148** |
| | HLA-B*38:01 | 12.5% | 3.6% | 0.0780 | 16.1% | 4.8% | $1.35 \times 10^{-3}$ |
| | HLA-C*07:01 | 20.8% | 14.4% | 0.283 | 29.0% | 12.6% | $8.380 \times 10^{-4}$ |
| | HLA-C*12:03 | 20.8% | 14.9% | 0.304 | 32.3% | 12.0% | $5.530 \times 10^{-5}$ |
| | HLA-A*25:01 | 4.2% | 5.9% | 0.772 | 14.5% | 4.2% | $2.079 \times 10^{-3}$ |
|  | HLA-B*18:01 | 8.3% | 11.7% | 0.790 | 16.1% | 6.9% | **0.0134** |

One-sided Fisher’s exact test was used to identify HLA-A, HLA-B and HLA-C alleles overrepresented in each of RS groups in cohorts of deceased and control individuals. Data shown for alleles with $p < 0.05$ in at least one of cohorts and with frequency over 5% in at least one of four considered conditions.

Finally, we assessed the contribution of individual peptides from different viral proteins to the RS. Contribution of a peptide to RS was calculated as an absolute value of the sum of corresponding PC coefficients (PC2, PC3 for HLA-A and PC4 for HLA-C). Then, the set of the most RS contributing peptides was composed by taking top 5% peptides from the corresponding distribution. The results of the procedure are summarized in Table 3: distribution of peptides with the strongest contributions over SARS-CoV-2 proteins was similar to the one calculated for all peptides after multiple testing correction. Without the correction, only the nonstructural protein 8 (NSP8) had a statistically significant odds ratio. Thus, considered peptides were spread over proteins without any significant dependence on contribution to the RS.

**Table 3.** Distribution of peptides and viral proteins and their contribution to the risk score (RS).

| Protein | Percentage,<br>important peptides | Percentage,<br>all peptides | Odds<br>ratio | *p* | FDR |
| --- | --- | --- | --- | --- | --- |
| NSP3 | 19.5% | 19.6% | 1.0 | 1.000 | 1.000 |
| Spike | 9.5% | 12.2% | 0.8 | 0.164 | 0.682 |
| NSP12 | 13.7% | 10.4% | 1.4 | 0.0647 | 0.539 |
| NSP4 | 5.2% | 7.2% | 0.7 | 0.186 | 0.582 |
| NSP13 | 5.5% | 6.2% | 0.9 | 0.724 | 0.953 |
| NSP2 | 6.1% | 5.3% | 1.2 | 0.528 | 0.825 |
| NSP14 | 4.0% | 5.2% | 0.8 | 0.439 | 0.784 |
| NSP6 | 3.7% | 4.7% | 0.8 | 0.423 | 0.814 |
| NS3 | 2.1% | 3.5% | 0.6 | 0.215 | 0.597 |
| NSP16 | 4.3% | 3.3% | 1.3 | 0.339 | 0.770 |
| M | 5.2% | 3.1% | 1.7 | 0.0532 | 0.665 |
| N | 3.7% | 3.0% | 1.2 | 0.508 | 0.847 |
| NSP5 | 2.4% | 2.9% | 0.8 | 0.737 | 0.921 |
| NSP15 | 4.3% | 2.6% | 1.7 | 0.0800 | 0.500 |
| NSP8 | 4.3% | 2.0% | 2.2 | **$9.84 \times 10^{-3}$** | 0.246 |
| NS7a | 1.8% | 1.5% | 1.2 | 0.640 | 0.889 |
| NSP1 | 1.5% | 1.4% | 1.1 | 0.806 | 0.959 |
| E | 0.9% | 1.2% | 0.8 | 1.000 | 1.000 |
| NS8 | 0.3% | 1.2% | 0.3 | 0.272 | 0.679 |
| NSP9 | 0.6% | 1.1% | 0.6 | 0.584 | 0.859 |
| NSP7 | 0.0% | 0.8% | 0.0 | 0.178 | 0.637 |
| NS6 | 0.3% | 0.6% | 0.5 | 1.000 | 1.000 |
| NS7b | 0.0% | 0.5% | 0.0 | 0.405 | 0.845 |
| NSP10 | 1.2% | 0.5% | 2.6 | 0.083 | 0.417 |
| NSP11 | 0.0% | 0.0% | 0.0 | 1.000 | 1.000 |

Fisher’s exact test was used to compare fractions of each protein in the most RS contributing and all peptides.

### HLA class I homozygosity is a double-edged sword for COVID-19 risk

When analyzing the high risk score group we noticed that more than a half (five out of eight) deceased patients containing HLA-A*01:01 alleles were homozygous by this allele while the medium group had not a single individual homozygous by HLA-A*01:01 (Fisher’s exact test *p* = 0.0103). The distribution of individuals homozygous by HLA-A*01:01 among the groups of deceased patients and the control group proves the negative role. It turned out that the distribution in the deceased group (4 out of 26 patient who died at the age not greater than 60 and 1 out of 85 patients who died at the age greater than 60) leads to two statistically significant differences: *p*-value of Fisher’s exact test comparing the adults group with the elders group equals 3.10×10^−3^, and *p*-value of the test comparing the adults group and the control group equals 0.0104 (8 out of 428 members of the control group were homozygous by HLA-A*01:01). However, the difference between the elders group and the control group is statistically insignificant (*p* = 0.155). Interestingly, there were no other statistically significant differences in the distribution of homozygosity between the groups.

Generally, the average age of death for patients homozygous by any allele was significantly less compared to heterozygous ones (Mann-Whitney U test *p* = 6.45 × 10^−3^, S2 Fig). Also the fraction of homozygous patients was higher in the group of deceased adults (42.3%) compared both to elderly patients (15.3%, Fisher’s exact test *p* = 6.03×10^−3^) and the control group (19.2%, *p* = 9.80×10^−3^). Difference between elderly and control groups was not statistically significant (*p* = 0.448).

However, the low risk group also contained homozygous individuals: all homozygosity cases by HLA-A*02:01 (six cases) and HLA-A*03:01 (two cases) alleles were associated with low risk. These assignments were in agreement with age of death: only one patient homozygous by HLA-A*02:01 had not passed the 60 years age of death threshold. Thus, homozygosity by HLA class I genes is generally associated with poor prognosis except some alleles like HLA-A*02:01 and HLA-A*03:01 with “relevant” peptide-binding profiles.

## Discussion

In the current study, we presented the characteristics of a large cohort of deceased patients with COVID-19 from O.M. Filatov City Clinical Hospital in Moscow, Russia. The clinical characteristics of these patients indicated that the HLA genotype, age and underlying diseases were the most important risk factors for death. In our population the median age of deceased patients was 73.0 years. Three previous studies reported an average age in non-survivors respectively of 78.0, 65.8 and 70.7 years-old [24–26]. Our data are in line with the literature reaffirming that advanced age is one of the strongest predictors of death in patients with SARS-CoV-2 [27].

The majority of deceased patients at age not greater than 60 were men (61.5%), while the populational level for this age category in Russia is 48% [28]. Such imbalance is in agreement with information that COVID-19 is more prevalent in men [27]. This trend continued on group of elderly adults where sex distribution was close to uniform, while only 37.5% of population from this age group are males.

Only 18 deceased elderly patients (21.2%) had not any comorbidities. A number of previous studies mentioned high percentage of comorbidities in group of patients with severe course of COVID-19 [24, 28, 29]. At the same time, we were unable to find statistically significant differences in fractions on different comorbidities between adult and elderly group except the percentage of cerebrovascular disease. Specifically, only one adult (3.8%) had suffered from this disease compared to 29 individuals (34.1%) from the older group. The results of previously conducted meta-analysis of 1558 individuals infected by COVID-19 already highlighted cerebrovascular disease as a risk factor for COVID-19 infection, however this was not associated with increased mortality [30]. It is well known that frequency of cardiovascular comorbidities increases with age [31], and in total with age-related decrease in T-cell receptor repertoires it negatively affects prognosis of COVID-19 [32].

Differences of frequencies of HLA class I alleles were not statistically significant after multiple testing correction both in comparisons between deceased patients and control group, and deaths of adults versus elderly ones. Previously, Wang with co-authors performed comparisons of allele frequencies between groups of Chinese COVID-19 infected individuals with control ones which resulted in significant difference only for rare alleles such as HLA-C*07:29 and HLA-B*15:27 [33]. Thus, the analysis of the whole HLA class I genotype should be performed to identify possible associations with clinical information.

Since the available cohort size is insufficient to deeply cover possible genotypes (two alleles for each of HLA-A, HLA-B and HLA-C genes) we assigned a numerical value to each allele associated with aggregate binding affinity of viral peptides to the corresponding receptor. The obtained risk score (RS) separated adult patients died due to COVID-19 both from elderly ones and the control group with a statistical significance. A conceptually similar technique was used by Iturrieta-Zuazo et al: allele score was calculated as a number of tightly binding viral peptides (affinity less than 50 nM) [10]. However, our PC-based approach can be more robust since it does not depend on any threshold.

To identify extreme values of RS we splitted its range into low, medium and high risk groups. Three HLA-A alleles were highly overrepresented in these groups: HLA-A*02:01 and HLA-A*03:01 were tightly associated with low risk while HLA-A*01:01 contributed to the high risk group. Connection of HLA-A*01:01 and HLA-A*02:01 alleles with COVID-19 morbidity and mortality was already mentioned in the existing literature. Namely, frequency of HLA-A*01:01 in Italian regions positively correlated both with number of COVID-19 cases and deaths, while significant negative correlation was observed for HLA-A*02:01 [11].

An interesting illustration for the role of HLA-A*01:01 and HLA-A*02:01/HLA-A*03.01 was discovered during the analysis of homozygous individuals. Homozygosity only by HLA-A*01:01 as well as homozygosity by any allele were significantly associated with earlier age of death compared to the corresponding heterozygous individuals. Such observation was already noted for some other infectious diseases. For example, limited number of recognized peptides due to HLA class I homozygosity lead to higher progression rate from HIV to AIDS [34]. On the contrary, we found that only one out of eight HLA-A*02:01 or HLA-A*03:01 homozygous individuals died before 60 years. This fact can be also observed in dataset recently published by Warren with co-authors [35]: none out of five COVID-19 patients homozygous by HLA-A*02:01 or HLA-A*03:01 had severe course of COVID-19 and were admitted to intensive care unit. Thus, presentation of “important” viral peptides with doubled intensity can enhance immune response showing that HLA class I homozygosity can act like a double-edged sword.

Since RS was constructed as a linear combination of peptide-HLA binding affinities, it was possible to rank peptides according to their contribution to the RS. Only one protein, NSP8, had a statistically significant fraction of RS contributing peptides, which, however, was negligible after multiple testing correction. Thus, the most “important” peptides were spread across viral proteins proportional to their total fractions (see Table 3). These results suggest that spike protein-based vaccines at phase 3 clinical trials (Gamaleya Research Institute, BioNTech/Fosun Pharma/Pfizer, Moderna/NIAID, University of Oxford/AstraZeneca, CanSino Biological Inc./Beijing Institute of Biotechnology, Janssen Pharmaceutical Companies and Novavax) can be no worse than inactivated vaccines in terms of HLA class I presentation.

## Data Availability

All relevant data are within the manuscript and its Supporting Information files.

## Data availability

All relevant data are within the manuscript and its Supporting Information files.

## Supporting information

**S1 Fig. Distribution of HLA-A, HLA-B and HLA-C alleles in risk score (RS) groups**. Alleles with frequency over 5% in the cohort of deceased patients or in the control group are presented.

**S2 Fig. Age at death of homozygous and heterozygous deceased COVID-19 patients**.

**S1 Table. HLA class I genotypes of deceased patients and the control group**.

**S2 Table. GISAID identificators of used SARS-CoV-2 genomes**.

**S3 Table. Raw and processed HLA-peptide binding affinity matrices**.

**S4 Table. Statistical comparison of allele frequencies in deceased adults, elderly adults and the control group**.

**S5 Table. Percentage of the explained variance for HLA-A, HLA-B and HLA-C principal components**.

**S6 Table. Comparison of principal components and risk score between deceased adults, elderly adults and the control group**.

**S7 Table. Distribution of deceased adults, elderly adults and the control group in low, medium and high risk score groups**.

