## Supplementary figures and images for "Association of HLA class I genotypes with age at death of COVID-19 patients"

### S1 Fig

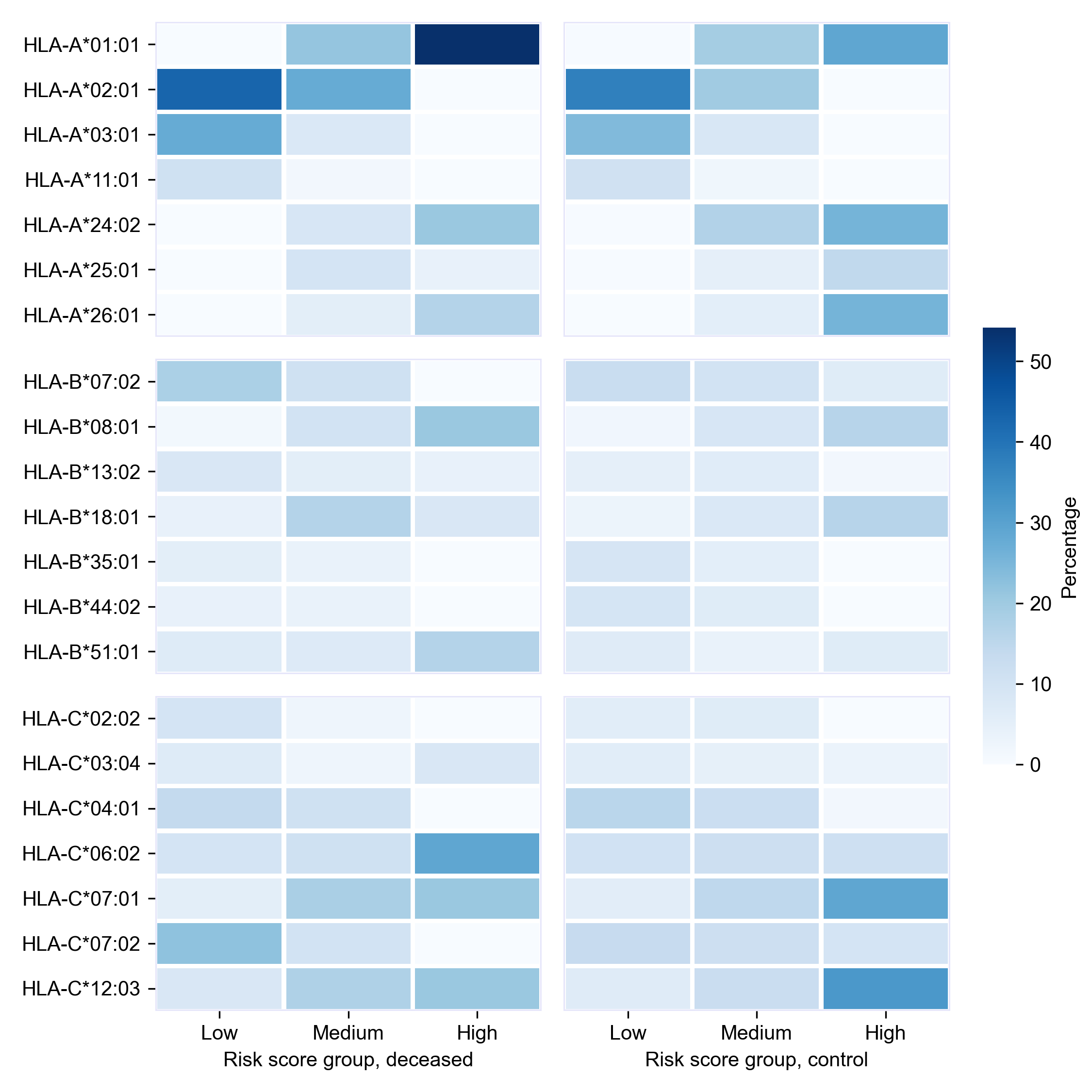

### S2 Fig

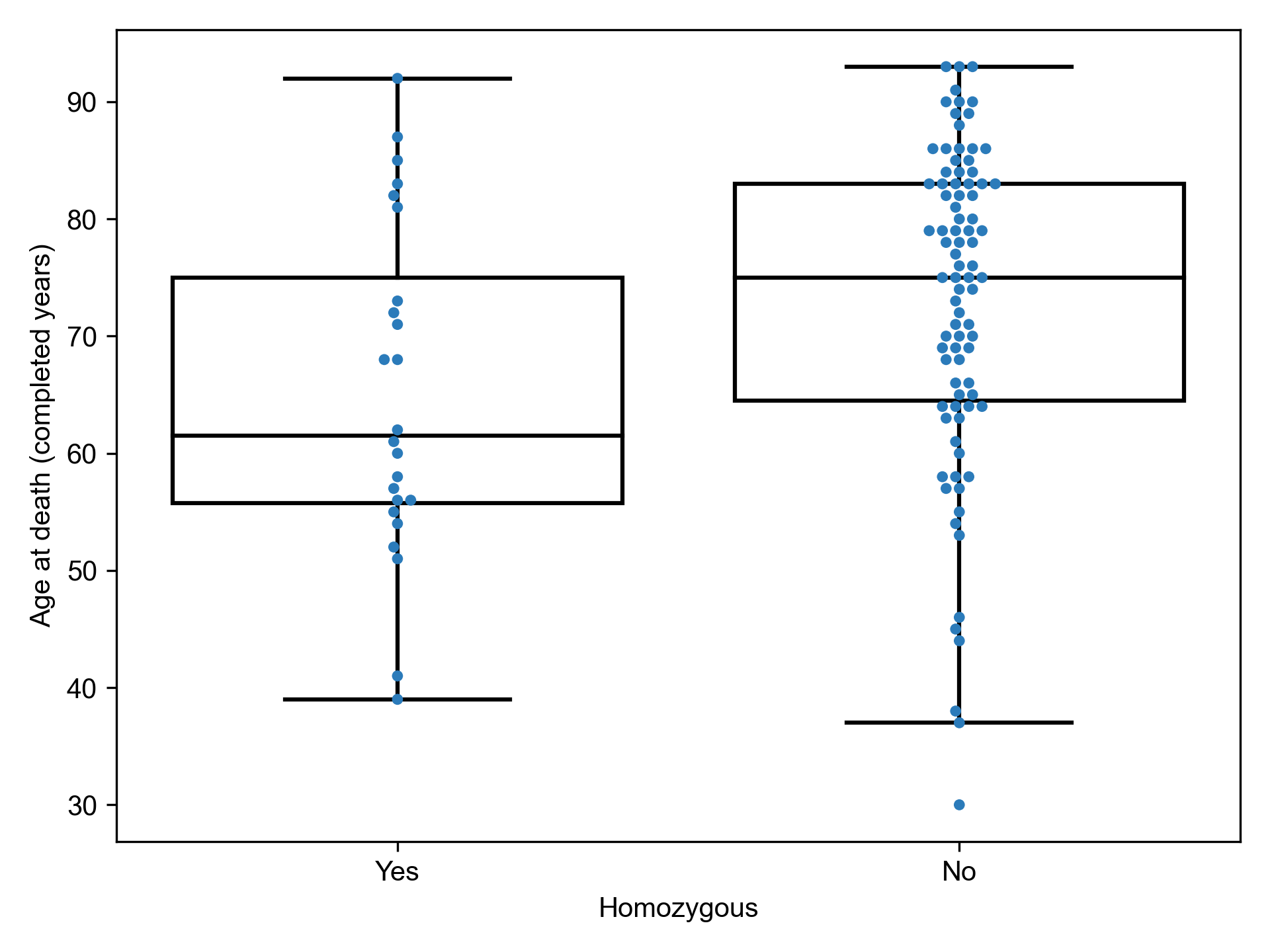
